# Comparing Modelling Architectures in the context of EGFR Status Classification in Non Small Cell Lung Cancer

**DOI:** 10.64898/2026.02.16.26346059

**Authors:** Owen Anderson, Russell Hung, Simon Fisher, Alexander Weir, Jeremy P. Voisey

## Abstract

Radiogenomics enables the non-invasive characterisation of the genomic and molecular properties of tumours, with epidermal growth factor receptor (EGFR) mutations in non–small cell lung cancer (NSCLC) being one of the most investigated applications. In this study, we evaluate radiomics, contrastive learning, and convolutional deep learning approaches to predict the EGFR mutation status from chest Computed Tomography (CT) images using the TCIA Radiogenomics dataset (n=115). Our results, using 10-fold cross validation, demonstrate the capacity of imaging models to predict mutation status from CT data in a manner consistent with existing literature. Among the evaluated methods, models integrating radiomic with clinical features achieved the best performance, with an AUC of 0.790 and AUPRC of 0.517, outperforming both contrastive learning (AUC=0.787) and convolutional architectures (AUC=0.763). Beyond methodological comparisons, we discuss the challenges related to clinical translation. Specifically, we contrast radiogenomics with conventional tissue biopsies, and identify scenarios where radiogenomics might be useful, either independently or in conjunction with other existing diagnostic technologies. Together these findings evidence the potential utility of radiogenomics EGFR models and provide direct architecture comparisons on the same dataset.

## Introduction

Lung cancer contributes to more deaths than any other type of cancer and is expected to increase in global prevalence (1, 2). Recent advances in precision medicine have facilitated the stratification of lung cancer patients into subgroups based on the presence of actionable mutations, defined as somatic variants which can be treated with targeted-agents. Actionable mutations typically occur within protein-coding regions of genes that mediate oncogene addiction, rendering them susceptible to pharmacological inhibition (3, 4). In non-small cell lung cancer (NSCLC), kinase-domain exonic variants of the epidermal-growth-factor-receptor (EGFR) gene represent well-characterised actionable mutations. EGFR genotyping facilitates the stratification of patients for treatment with tyrosine kinase inhibitors (TKIs), which have been shown to improve survival outcome and reduce recurrence rates for EGFR+ NSCLC (5–7).

The genotyping of NSCLC conventionally depends on direct tissue sampling of the target lesion (8). This is readily available for patients who have undergone surgery. For non-surgical patients either transbronchial (following the airway) or transthoracic (penetrating the chest wall) biopsies are performed. However, patient and disease phenotypes such as co-morbidities, frailty and inaccessible lesion locations can be detrimental to biopsy success likelihood and may lead to complications. Among those who undergo lung biopsies, pneumothorax and internal hemorrhage are some possible side-effects (9, 10). Additionally, failure rates for such procedures can be as high as 20% for sample collection and close to 50% for next-generation sequencing when there is insufficient tissue (11–13). In such cases, patients would either need to repeat the invasive procedure or progress to standard systemic anti-cancer therapy (SACT) due to the absence of useful genotyping information. There might be the potential for *radiogenomics* to aid diagnostic decisions in these cases.

We define radiogenomics as the non-invasive classification of somatic genetic variants using medical imaging modalities. Radiogenomics is performed on lung Computed Tomography (CT) scans with two main methods. Firstly, radiomics, the quantitative extraction of features from medical images to describe patterns such as texture, shape, and intensity, which are fed into classification algorithms (14). The other method applies convolutional deep learning to CT images, either directly classifying the mutation status or towards producing representations that can be used by downstream classification algorithms.

The classification performance of radiogenomic models is crucial for clinical adoption. In pre-existing literature, radiomic models that classify EGFR status in NSCLC achieved AUC performance with a range of 0.64–0.95 (mean 0.77) on the validation set. Studies generally adopt a pipeline of feature extraction and selection followed by algorithm fitting. Machine learning algorithms, such as logistic regression, random forest and support vector machine, are common. Training set size in radiomic models ranges from fifty patients, to several hundred (15–30). In contrast, deep learning models have a tighter spread with a validation AUC performance range of 0.71–0.84 (mean 0.82) and employ 2D or 3D residual neural networks of varying depth and complexity. Dataset sizes in deep learning models are generally larger, with a few studies having up to several thousand patients (23, 25, 31–38). In general, the results from literature suggest that integration with other modalities, particularly clinical data, and sometimes deep learning features, boosts performance (23, 25, 33, 36, 37).

Whilst conventional radiomic and deep learning approaches are common for EGFR prediction, some methods allow for unsupervised learning and therefore the use of unlabelled data. Contrastive learning is one such method, whereby the model learns to represent the data by contrasting similar and dissimilar examples. Contrastive learning has been applied to digital pathology whole-slide imaging for mutation prediction, with positive results (39–41), but its use in CT imaging and radiology is rare. In this work we explore its use to produce useful representations for EGFR status classification. In particular we look at an implementation of contrastive learning known as the SimCLR framework, which was first described by Chen et al in 2020 (42).

Using the publicly available NSCLC Radiogenomics cohort (43), we establish baseline performance of both radiomics and deep learning models, both with and without integration with clinical features. Subsequently, we compare these baseline models with representations generated by SimCLR models.

## Methods

### Dataset and Processing

The NSCLC Radiogenomics dataset (43) was used in all experiments. This dataset consists of CT, semantic annotations of tumours, paired gene mutations and clinical data from a cohort of NSCLC patients (n=211). Of the entire dataset, the number of samples for which a segmentation mask and an EGFR genotype were both present was 115. The original DICOM CT scans were sorted in anatomical sequence, cropped to maintain consistent spacing, and converted to NIfTI-2 format. All volumes were resampled to isotropic 1 mm spacing and windowed with a lung window (window level -600 HU, window width 1500 HU) using the ITK framework (44). For clinical data, one-hot encoding was applied to categorical data, continuous features were standardised, and K-nearest neighbours imputation was used to impute missing features. A total of 7 clinical features were used in the experiments. The 115 cases were split into 10 folds stratified on the EFGR label. Identical fold assignments were maintained across experiments to facilitate model comparison.

### Radiomics Extraction

The Pyradiomics library (14) was used to generate radiomic features from the volumes using the specific configuration from their example documentation with bin width set to 20. Radiomic features were generated separately from a 9 mm dilation created around the segmentation mask to create peritumoral features; the peritumour was considered a separate entity from the tumour during radiomic feature generation. In total, 200 radiomic features were generated from each region of interest (ROI) — 100 from the tumour region and 100 from the peritumoral region.

### Generation of Contrastive Features

Contrastive learning aims to minimise the distance in latent space between similar instances and maximise it between dissimilar samples. We adapt the SimCLR framework (42) (Figure 1) to generate representations from CT images. The network is composed of a 2D ResNet-18 followed by a multi-layer perceptron to generate a latent vector. The SimCLR loss is used (45). Firstly, the framework is trained on ImageNet^i^, where each training batch is constructed to include augmented examples of the same image (similar), which are contrasted with other (dissimilar) images in the dataset. Secondly, this model is further fine-tuned on image pairs taken from the LIDC-IDRI (46) and NSCLC-Radiomics (47) datasets. We expect many characteristics (including mutational statuses and biological processes) to be more similar when sampling spatially distinct regions within the same tumour compared to sampling regions from different tumours. As such, similar pairs are taken from different sections of the same tumour. These images are then masked to a ROI, which removes irrelevant information from the images. The ROI is generated by taking the existing manually generated segmentation for each image, and dilating this segmentation by 5 voxels to include the peritumoral region, then masking the CT image to this region. Flipping was randomly applied between the images in each pair.

**Fig. 1.**
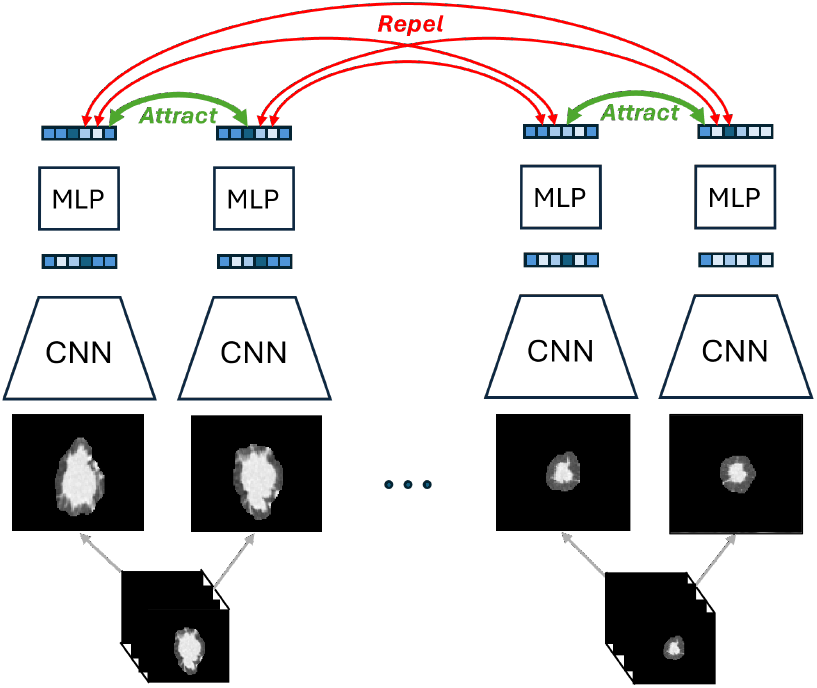
Contrastive learning approach: The CNN and MLP components are jointly optimized to produce representations. The contrastive approach ensures that slices from the same tumour have a similar representation, and slices from different tumours are encouraged to be dissimilar. Through this training process, the model learns semantically meaningful representations.

### Machine Learning

Features generated by either Pyradiomics or contrastive learning models were used to train logistic regression models to predict the EGFR mutation status. All features were standardised, and feature selection was performed using analysis of variance (ANOVA) with a significance threshold of p < 0.5. L1 (lasso) regularisation was used. Hyperparameters were optimised using grid search within a cross-validation framework. Two strategies for integration of clinical and radiomic/contrastive features were compared. In early integration, clinical and radiomic/contrastive features were concatenated before being fed into a logistic regression model, while late integration uses the product of the predicted probabilities of individual clinical and radiomic/contrastive classifiers.

### Deep learning

We explored both 2D and 3D deep learning approaches. For 2D, the image slice with the largest ROI area was used to train a convolutional neural network to predict EGFR status. The 2D CNN consisted of 3 convolutional layers then max pooling followed by two fully-connected layers. For 3D, a ResNet18 was used to predict the EGFR status from the image patches. Image patches of 50 × 50 × 50 voxels (1 mm isotropic) were extracted from the original volume centred on the centroid of the segmentation mask. For models where clinical features were integrated with image patches, standardized clinical features were concatenated with the image representation prior to the penultimate fully connected layer of the neural network (Figure 2).

**Fig. 2.**
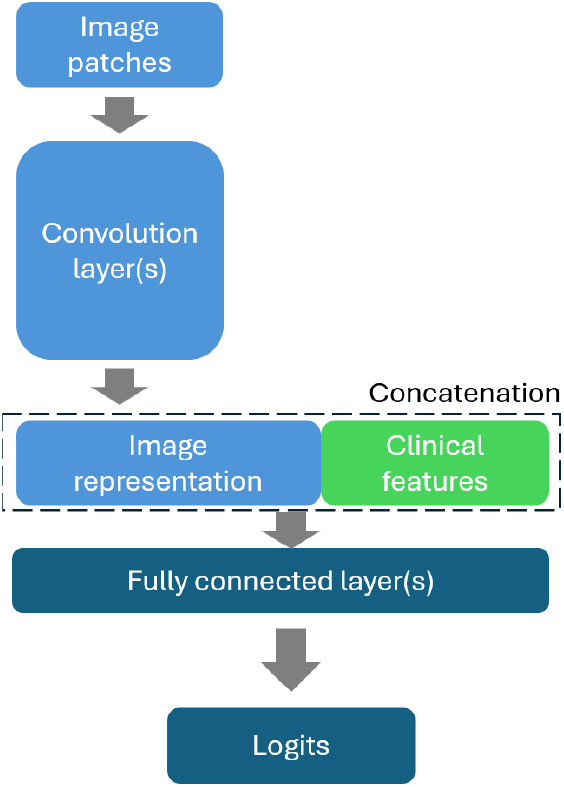
Schematic diagram of integrating clinical features in deep learning training.

**Fig. 3.**
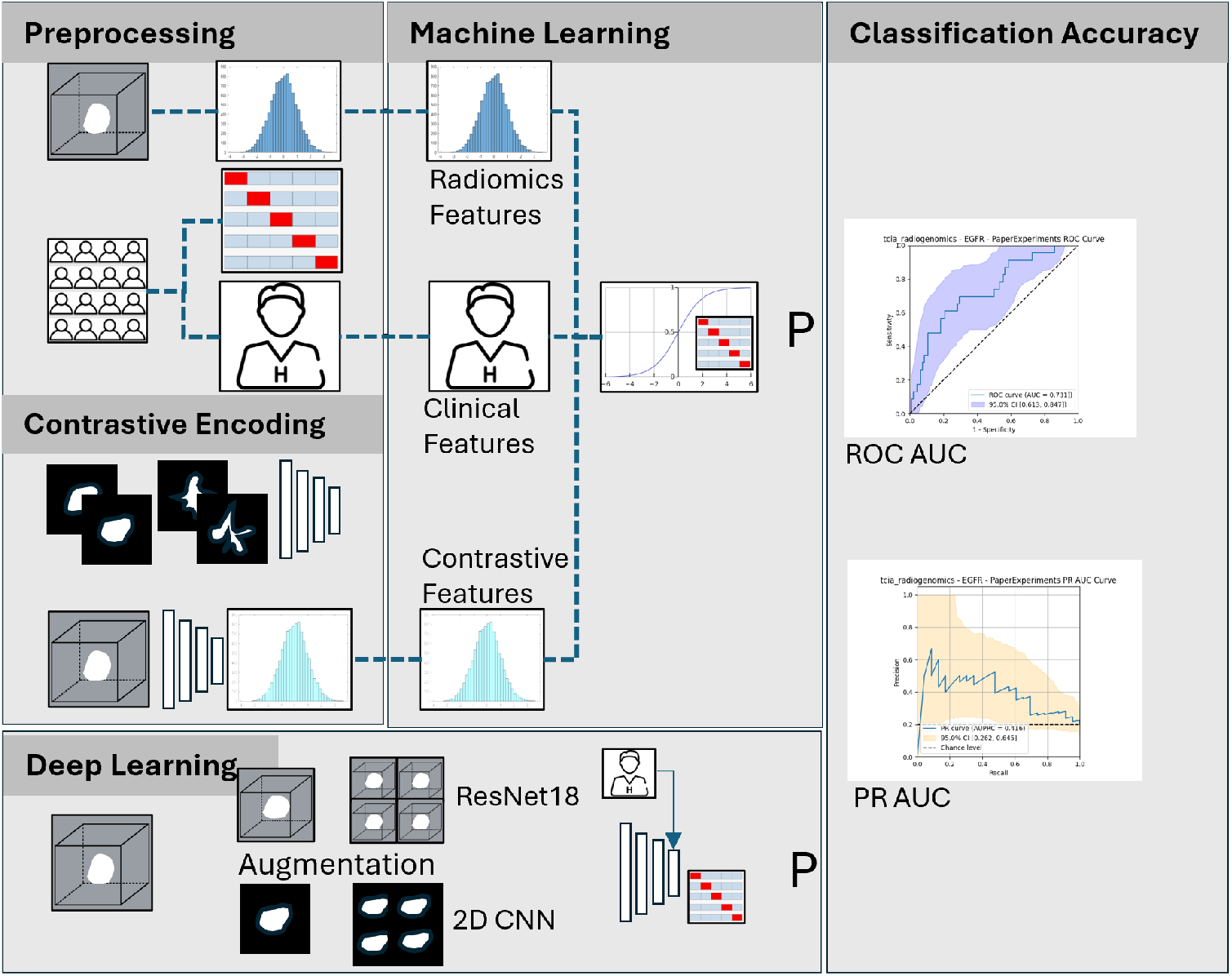
Graphical representation of the methods used within the paper. For preprocessing, radiomic features were extracted from the volume whilst clinical and 10-fold splits were generated from the clinical data. A contrastive loss model was either frozen as pretrained on ImageNet or fitted to nodule pairs derived from the lung tumour images, and the intermediate layer of this used to produce “contrastive encodings”. Both 2D and 3D deep learning approaches were used with augmentation prior to fitting. The logits or probabilities of resulting fitted models were used in out-of-fold evaluation for model classification accuracy with both AUC and AUPRC.

Image augmentation was implemented by applying a series of transforms to the original image. These transforms include flipping, rotation, and scaling in all dimensions. In the 2D case, we also apply translation and randomness in the selection of the image slice. A random slice was picked where the area of the ROI was higher than or equal to 70% that of the largest slice.

### Model Evaluation

Following the recommendations for assessing the performance of predictive models we use discrimination, calibration, and decision-analytic measures (48). All models were iteratively fitted on the training folds and predictions generated using the validation fold using the splits previously described. Performance was then evaluated using the out-of-fold predictions. To measure discrimination we used the area under the receiver operating characteristic (AUC) and precision recall curves (AUPRC). For these, bootstrapping using 1000 iterations was used to calculate the 95% confidence intervals.

## Results

### Machine Learning with Radiomic Features

Using only clinical features resulted in a performance of AUC=0.760 and was superior to both the radiomics model at 0.667 and the early integration model at 0.731. However, the late integration model using the product of the prediction for both clinical and radiomic models was superior to all models at 0.790 (Table 1, Figure 4). Evaluating using AUPRC, the clinical-only model had an AUPRC of 0.384 and the radiomics model 0.305. For this metric, early integration outperformed clinical with AUPRC=0.389, but was itself outperformed with the late integration model at 0.517. For both metrics, the late integration model combining clinical and radiomics was the best performing.

**Table 1.**
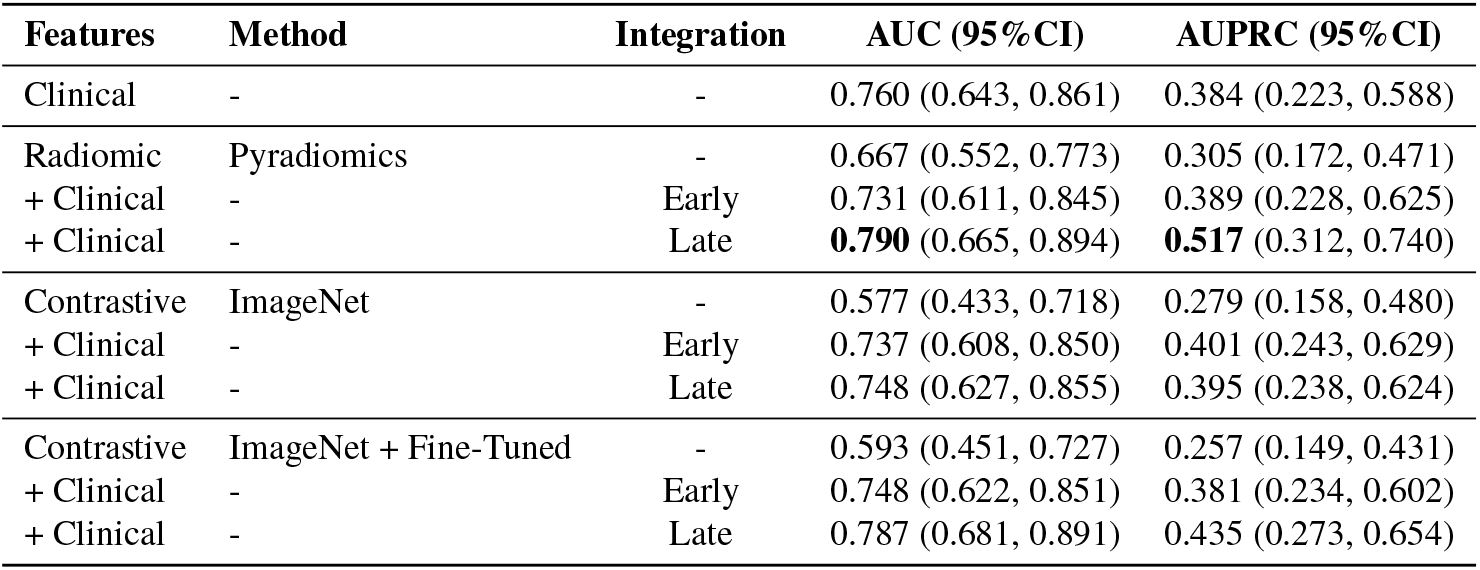
10-fold cross validation out-of-fold performances of TCIA Radiogenomics models (n=115). The performance of logistic regression models applied to radiomic features and to features derived from contrastive learning is presented. Contrastive representations were extracted from models only trained on ImageNet as well as from models further fine-tuned on lung imaging data. Models based solely on radiomic or contrastive features were evaluated both independently and in combination with clinical features. A model using only clinical variables was included as a control for comparison. Results shown as value (95%CI) using bootstrapped predictions (1000 iterations).

**Fig. 4.**
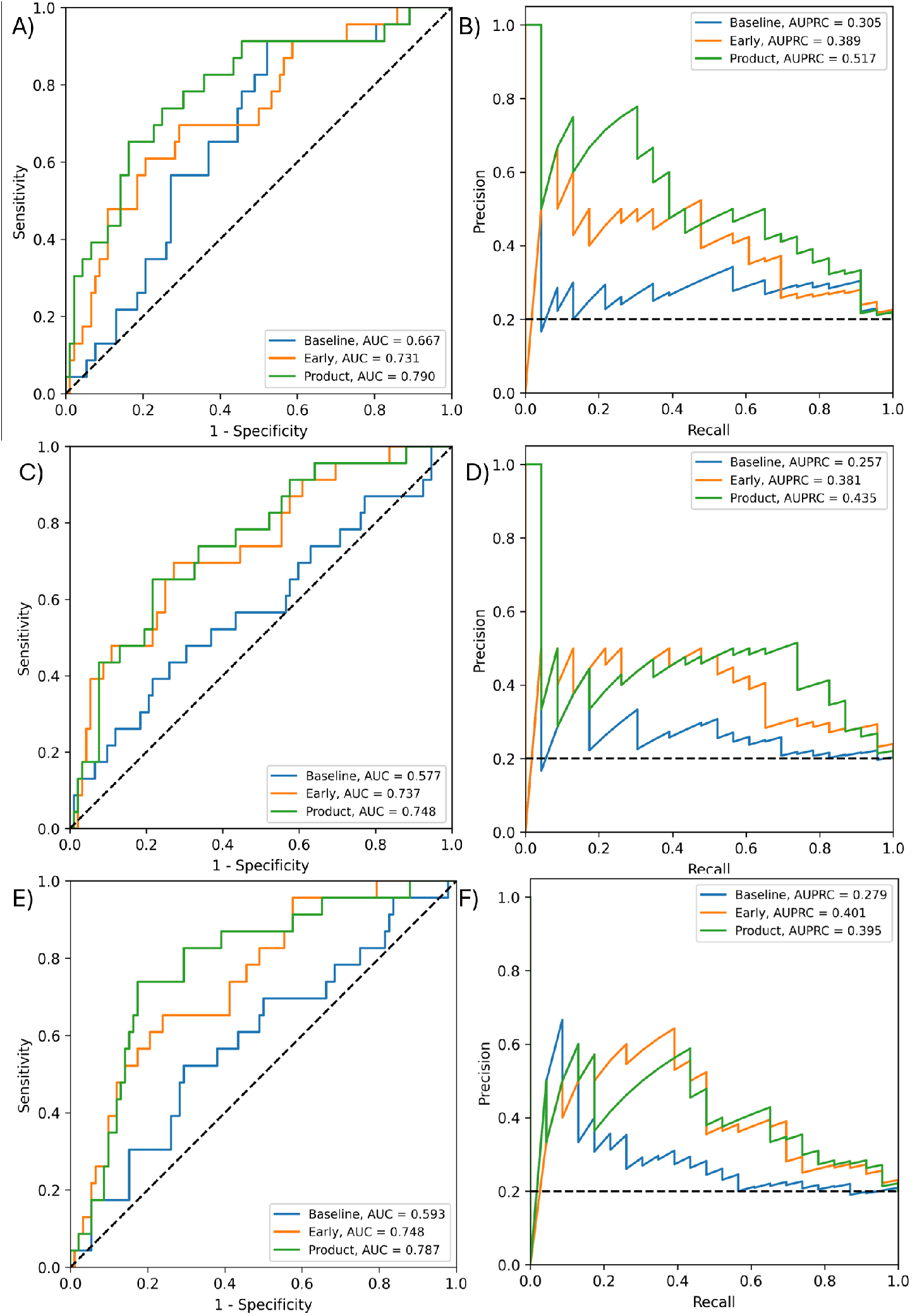
Out-of-fold performance of logistic regression models using radiomic features (A & B) and embeddings generated by contrastive learning without fine tuning (C & D) and with fine tuning (E & F). ROC (A, C, E) and precision-recall (B, D, F) curves for logistic regression models fitted using radiomic features, ImageNet contrastive features and fine-tuned contrastive features respectively. Baseline (blue) refers to models without clinical features. Models that integrate clinical features using the early strategy are shown in orange, and those using the late (product) strategy are shown in green.

### Machine Learning with Contrastive Loss Features

Contrastive representations were obtained from two contrastive models: 1) trained on ImageNet only, 2) trained on ImageNet and fine-tuned with lung imaging data from the LIDC-IDRI and NSCLC-Radiomics dataset. Table 1 shows the results of fitting logistic regression models fitted on the contrastive representations, either on their own or together with clinical features. Fine-tuning on lung imaging data improved the AUC score (0.593 vs. 0.577). As expected, the integration of clinical data improved performance in both cases, with late integration outperforming early integration. Figure 4.

### Deep Learning

The 2D deep learning with clinical data and augmentation was the best performing baseline deep learning model in terms of AUC (0.763). The addition of clinical features resulted in a reduction in AUPRC, regardless of whether augmentation was used. Using augmentation consistently improved AUC and AUPRC for the 2D CNN. Using a 3D ResNet18 did not outperform the 2D CNN. The ResNet18 with the best performance included clinical features but without augmentation (AUC=0.754). Adding clinical features improved performance of 3D ResNet in general (Table 2, Figure 5).

**Table 2.** 10-fold cross validation out-of-fold performances of TCIA Radiogenomics deep learning models (2D CNN or Resnet18) (n=115). Clinical features were added prior to the penultimate layer of each classifier architecture.Results shown as value (95%CI) using bootstrapped predictions (1000 iterations).

| Model | Clinical | Augmentation | AUC (95% CI) | AUPRCC (95% CI) |
| --- | --- | --- | --- | --- |
| 2D CNN | No | No | 0.713 (0.587, 0.837) | 0.424 (0.242, 0.633) |
| 2D CNN | No | Yes | 0.720 (0.609, 0.825) | **0.464 (0.270, 0.630)** |
| 2D CNN | Yes | No | 0.735 (0.618, 0.837) | 0.385 (0.229, 0.584) |
| 2D CNN | Yes | Yes | **0.763 (0.652, 0.861)** | 0.395 (0.238, 0.595) |
| ResNet18 | No | No | 0.596 (0.470, 0.725) | 0.235 (0.141, 0.355) |
| ResNet18 | No | Yes | 0.634 (0.506, 0.755) | 0.311 (0.169, 0.499) |
| ResNet18 | Yes | No | 0.754 (0.646, 0.851) | 0.397 (0.237, 0.599) |
| ResNet18 | Yes | Yes | 0.743 (0.642, 0.843) | 0.393 (0.223, 0.589) |

**Fig. 5.**
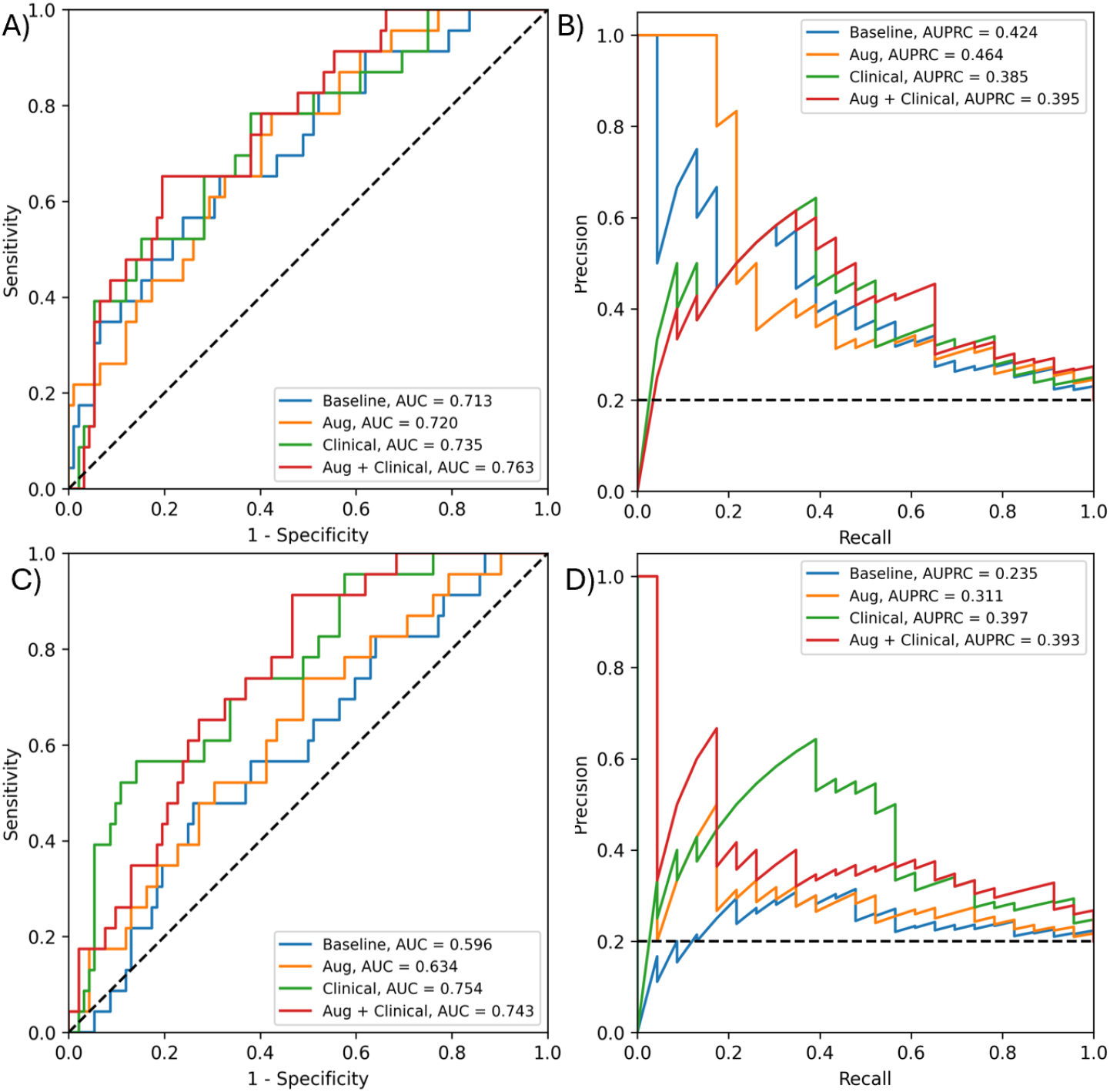
Performance of 2D CNN (A & B) and ResNet18 (C & D) deep learning models. ROC (A) and precision-recall (B) curves for 2D CNN models in both the presence and absence of image augmentation and clinical features, generated from out of fold predictions using 10-fold cross validation. Clinical features were concatenated to the penultimate fully connected layer of the network. No augmentation or clinical features (blue), augmentation only (orange), clinical features with no augmentation (green), both augmentation and clinical features (red). ROC (C) and precision-recalll (D) curves for Resnet18 models in both the presence and absence of image augmentation and clinical features.

### Best Performing Models

Of all models architectures assessed, the best performing from each approach of radiomics (AUC=0.790), contrastive learning (AUC=0.787) and deep learning (AUC-0.763) are compared using ROC and precision-recall curves in Figure 6.

**Fig. 6.**
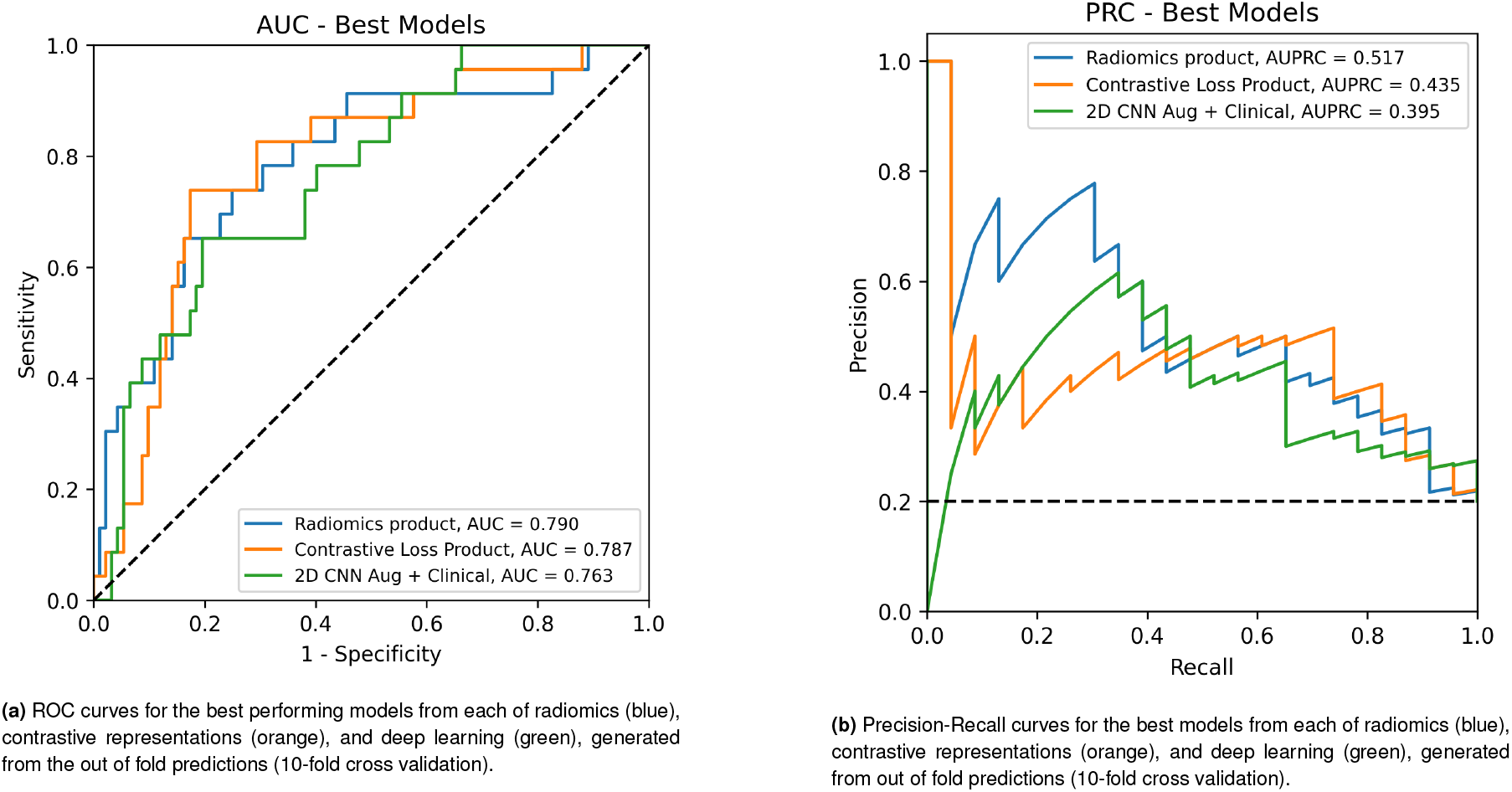
Key Results of top performing models as evaluated by AUC (a) and AUPRC (b)

## Discussion

In this paper, we demonstrate direct comparisons between radiomics, deep learning and contrastive representation networks for EGFR status prediction using the publicly available TCIA Radiogenomics dataset. Our results suggest that the status of EGFR can be predicted with moderate performance of up to an of AUC 0.790 using cross validation on this cohort. We observed this maximum performance when integrating radiomic and clinical features with a late strategy, which narrowly out-performed early integration. Both the deep learning and contrastive representation architectures were weaker in performance with maximum AUCs of 0.763 and 0.787 respectively. All best performing models from each architecture benefitted from integration with clinical features. Notably, clinical features alone were predictive with an AUC of 0.760. Contrastive representation features when used with imaging alone were poorly predictive with pretraining only. This performance increased with fine-tuning from 0.577 to 0.593 but was still poor. Deep learning with 2D CNN architecture offered the best imaging-alone performance at 0.713. Due to the limited dataset size and lack of holdout test set, these findings are indicative, but should be treated with caution with regards to their potential generalisability.

Our findings for EGFR prediction within this paper, at around AUC=0.78 maximum performance, are placed centrally within the observations of previous work of radiomics AUC=0.64–0.95 (mean 0.77) and deep learning AUC 0.71–0.84 (mean 0.82). The most useful comparisons can be made between external work which specifically examined prediction using the TCIA Radiogenomics dataset. In this context, Shiri et al. strongly outperform our work, with an AUC of 0.88 from baseline CT experiments with radiomics features (49). We are unable to reproduce this performance despite using an almost identical framework. One difference between the approaches is that Shiri et al. used Matlab radiomics rather than Pyradiomics. Though not reported here, we explored Matlab generated features, and found no significant improvement to our results. Furthermore, Le et al. also fit models to the TCIA Radiogenomics dataset and in their case the baseline radiomics models they present are similar in performance to our radiomics models, with the exception of their Genetic Algorithm feature selection approach which achieved an AUC of 0.89 (17). Both these papers employ deep approaches to model optimisation (either hyperparamter of feature-selection based) which may be challenging to perform without overfitting to this limited dataset, as we will discuss in limitations. Finally, Lu et al. explored different radiomics extraction methods in the context of TCIA Radiogenomics and present similar performance to our experiments during model development, but much lower in hold-out validation at AUC 0.56 for pyradiomics (29). They use a heterogenous dataset for their validation cohort which includes samples from TCGA-LUAD and TCGA-LUSC. The drop in performance observed in a blended validation dataset should be noted as a concern for the generalisability of homogenous TCIA Radiogenomics fitted models.

For physical biopsy sampling (specifically where the biopsy was adequate), NICE via the UK National External Quality Assessment Service (NEQAS) pilot scheme for EGFR-TK, reports the sensitivity and specificity of polymerase-chain-reaction (PCR) based assays at 99%, 95%CI (94%, 100%) and 69%, 95%CI (60%, 77%), where the retrospective presence of an EGFR TKI mutation was based on the objective response to TKI therapy in the patient (NICE HTG316). This indicates that baseline biopsy-based testing is highly likely to detect the presence of an EGFR mutation, but is also somewhat disposed to generate false positives. With respect to radiogenomics, the diagnostic accuracy thresholds of computer-aided EGFR prediction are not currently understood, as the technology is not embedded in any healthcare frameworks. Even though a sensitivity of around 60% as generated by our models did not outperform physical biopsy, there are scenarios where such a tool might still be useful, such as when biopsy does not obtain sufficient samples, or patients might be too frail to tolerate a biopsy procedure. In long-term patient monitoring where repeat biopsy might be needed, a radiogenomics model might provide an alternative.

Another such case is where a radiogenomics model supports further diagnostic action rather than dictating intervention. We posit that they may contribute to patient triage where they could be used to identify “high confidence” candidates which should be immediately biopsied for verification and conversely to de-prioritise patients whose imaging strongly suggests a lack of EGFR mutation. Using predictive models to prioritise patients is currently in trial or investigation for several radiology applications alongside sepsis triage (50–54). Additionally, where treatment recommendation is concerned, other non-invasive modalities could have additive benefits over the use of imaging alone. For example, imaging might be paired with circulating tumour DNA (ctDNA) findings to improve diagnostic accuracy. A positive signals from both radiogenomics and liquid biopsy may be much more likely to indicate a true positive than either alone as previously demonstrated for lung cancer diagonisis and prostate cancer detection and monitoring (55, 56).

In this work, we used the publicly available TCIA Radiogenomics dataset, which constituted 115 complete samples for both training and validation. For classification tasks, this should be regarded as a limited dataset. Where features were generated prior to model fitting, such as for both radiomics and contrastive features, the feature space was considerably larger than the number of samples, which violated predetermined guidelines (57). As a result, statistical feature selection was used to minimise the feature count but this was itself a caveated approach as it was performed in a cross-validation loop meaning different features were selected per-fold. For deep learning image classification, a dataset size of 115 is considered to be small (58) and we observed limited ability for both the 3D Resnet and the 2D CNN to fit to the data. We attempted to overcome these limitations by training our contrastive feature encoder on an extended dataset, however, we were still ultimately limited during the prediction task. Fundamentally, it is highly likely that both statistical-ML and deep learning EGFR prediction algorithms would benefit from increased data availability both in terms of performance and the reliability of interpretation of that performance. Furthermore, in this series of experiments we did not have access to a hold-out test set of any kind and therefore we cannot comment on the generalisability of the models presented here. Based on these limitations, future work should seek to make use of additional public or private datasets in order to support model fitting and evaluation. Both the TCGA and CPTAC data resources may constitute useful additional data and could be blended into a single heterogenous dataset for this task. Additionally, 3-dimensional contrastive encoders may be able to learn better representations of images such that EGFR classification performance may improve versus 2-dimensional approaches.

To conclude, these experiments provide evidence that EGFR status can be predicted from imaging with reasonable accuracy, and that a direct comparison between architectures on a small dataset suggests radiomics in combination with clinical features may be the best approach. However, the authors emphasise caution in strong interpretation of these findings due to the limited size of the dataset.

## Data Availability

All data used in the study were obtained from The Cancer Imaging Archive, using the NSCLC Radiogenomics dataset: DOI:10.7937/K9/TCIA.2017.7hs4

https://www.cancerimagingarchive.net/collection/nsclc-radiogenomics/

## Acknowledgments

The results shown here are in whole based upon data generated by The Cancer Imaging Archive, using the NSCLC Radio-genomics dataset: DOI:10.7937/K9/TCIA.2017.7hs46erv

## Footnotes

i https://lightning.ai/docs/pytorch/stable/notebooks/course_U_ vA − DL/13 − contrastive − learning.html

